# Clinical evaluation of a natural language processing system for assisting structured diagnosis recording at the point of care: MiADE (Medical Information AI Data Extractor)

**DOI:** 10.1101/2025.09.09.25334920

**Authors:** Mairead McErlean, Jack Ross, Jonathan Kossoff, Maisarah Amran, James Brandreth, Leilei Zhu, Gary Philippo, Wai Keong Wong, Folkert W. Asselbergs, Richard J.B. Dobson, Yogini H Jani, Enrico Costanza, Anoop D. Shah

**Author notes:** Corresponding author Anoop D. Shah Address: UCL Institute of Health Informatics, 222 Euston Road, London, NW1 2DA.

## Abstract

**Background:** Structured recording of key information such as diagnoses is essential for safe, efficient patient care, but is currently done incompletely because it is time consuming for clinicians. We developed a natural language processing system called MiADE integrated with the Epic electronic health record to provide suggestions for structured diagnosis entries at the point of care.

**Objectives:** To evaluate the usability, usefulness and impact of MiADE, and identify recommendations for systems to improve point of care structured data recording.

**Methods:** Mixed methods evaluation of the implementation of MiADE, with surveys, interviews and observed outpatient consultations. The number of structured diagnoses recorded per outpatient encounter was compared before and after MiADE, and completeness of inpatient problem lists was evaluated using the billing diagnoses as a gold standard.

**Results:** 85 clinicians consented to the study and were provided access to MiADE, and 24 used MiADE to receive structured data suggestions during the study period. Baseline survey data and observations showed wide variation in structured data recording despite clinicians considering it to be important. Half of post-implementation survey respondents considered MiADE to be ‘very’ or ‘moderately’ useful. Among outpatient users there was a 36% increase in the number of diagnoses recorded per encounter, but no improvement was seen in the inpatient setting.

**Conclusions:** Natural language processing using MiADE has the potential to improve structured data recording, but further development and better clinician engagement are needed in order to maximise its impact.

**SUMMARY BOX:** *What is already known on this topic:*

- Despite the benefits of structured recording of healthcare data for individual care and research, much of the key information in electronic health records (such as diagnoses) is only recorded in free text
- Structured data entry in electronic health record systems can be time-consuming and cumbersome for clinicians

*What this study adds:*

- Embedding natural language processing within the electronic health record to suggest structured data entries was reported by clinicians to be useful, and increased the recording of outpatient diagnoses
- Clinician engagement was challenging, and overall usage of structured data remained suboptimal

*How this study might affect research, practice or policy:*

- Usability of electronic health record systems needs to improve to enable clinicians to record high quality data without impeding their workflow
- Natural language processing embedded within electronic health records may improve ease of use for data entry, but high-level clinician buy-in is also needed to influence professional documentation practice

## INTRODUCTION

Data recorded in electronic health record (EHR) systems is vital for safe, effective patient care [1–3] and structured recording of key information such as diagnoses is recommended in professional guidance [4]. However, much of the information in current EHRs is in unstructured clinical notes (free text) [5], partly because it can be onerous and time consuming for clinicians to enter structured data [6].

Previous work to improve structured data recording has shown that tools are more likely to be successful if fully embedded in the workflow [7]. Natural language processing (NLP) has been suggested as a possible solution, but there are a lack of studies which have evaluated its effectiveness in improving structured documentation in EHR.

We therefore developed, implemented and evaluated a system called MiADE (Medical information AI Data Extractor) in a major NHS hospital. MiADE embeds NLP within the EHR user interface to assist structured data entry. The aims of our study were to: (i) capture clinician views on data documentation processes, (ii) evaluate the usability, usefulness and impact of MiADE, and (iii) develop recommendations to maximise the effectiveness of such systems.

## METHODS

This was a mixed-method, single-centre before-and-after study of the feasibility of improving structured diagnosis recording using a novel point of care NLP system. The study protocol was approved by the South Central - Hampshire A Research Ethics Committee (23/SC/0221) and is registered on the ISRCTN registry (ISRCTN58300671) [8].

### Structured diagnosis recording in the EHR

The EHR system used at UCLH, Epic, enables clinicians to enter diagnoses either in free text or in structured problem or diagnosis lists. For structured entries, clinicians need to choose a SNOMED CT concept [9] for each diagnosis, which typically involves keyword searching or browsing the SNOMED CT hierarchy.

MiADE provides an alternative way to enter SNOMED CT concepts, by automatically suggesting relevant concepts and displaying them alongside the original text when saving a note, via the Epic ‘NoteReader’ interface (Figure 1). The clinician can then add the relevant entries with just a few clicks. If MiADE is available for a user, they will see a ‘NoteReader’ button on their screen, which they can use to decide whether to invoke MiADE when saving the note. MiADE is non-interruptive; clinicians can ignore it and continue their usual workflow if they wish.

**Figure 1:**
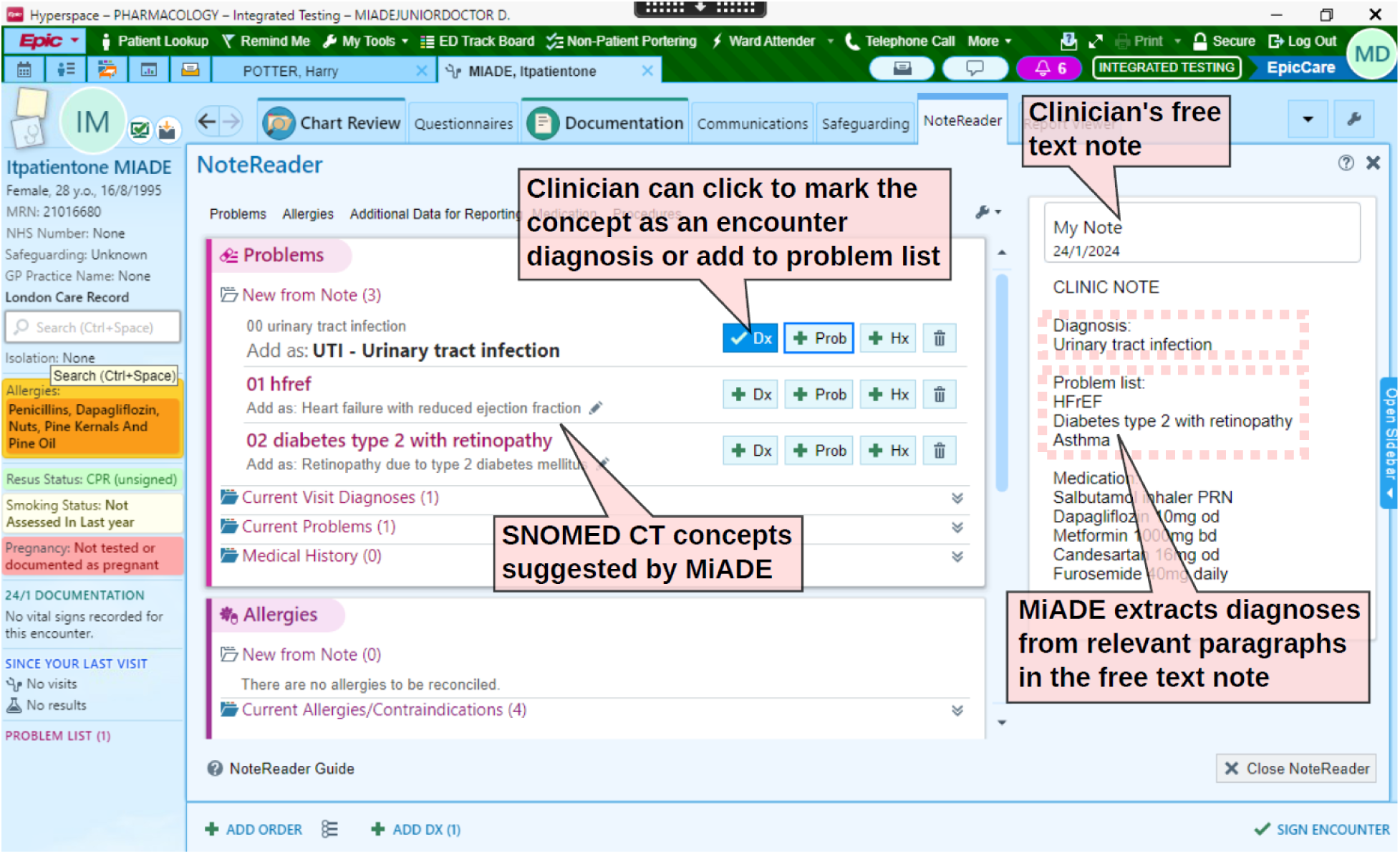
Screenshot of the Epic NoteReader electronic health record interface, which displays the clinical note and MiADE suggestions side by side. The clinician can add the SNOMED CT concepts suggested by MiADE to the problem list with a few clicks. The MiADE suggestions are prefixed by numbers to ensure that NoteReader displays them in the correct order. Copyright Epic Systems Corporation.

### MiADE design and implementation

MiADE was built using the MedCAT open-source NLP system [10] and underwent testing and Trust safety case approval before live implementation [11]. The evaluation study was advertised to clinicians by email, intranet blogs, and presentations at clinical meetings, and opened for recruitment on 27 December 2023. Participants were asked to watch a 5-minute training video and complete an online survey and consent form. A focus group was held one month after go-live, following which changes were made to the filtering of MiADE suggestions. MiADE was originally configured to return diagnoses and other clinical findings from paragraphs with a relevant header (e.g. ‘Problem list’) and up to one concept from elsewhere in the text. The change was to filter out non-diagnosis concepts (e.g. symptoms), and only analyse paragraphs with a relevant header.

### Study procedures

Individual clinician surveys were distributed before and after MiADE implementation using the Qualtrics XM platform, with a minimum two-week usage period required before completing the second survey. Observations of outpatient clinics were conducted in the same timeframe, aiming to observe at least five clinicians in both phases, with purposive sampling to include a range of specialties and seniorities (see Supplementary Figure). Post-implementation, at least five inpatient clinicians participated in one-to-one semi-structured interviews via MS Teams or telephone (see supplement for interview topic guide). Four MiADE investigators were hospital clinicians at the study site and were excluded from analysis.

### Qualitative data analysis

Audio transcripts of clinician and patient interviews were collected on a secure digital voice recorder and transcribed by a professional service. The interviews were 10-20 minutes in duration. The clinician workshop was recorded and transcribed via MS Teams. Observations and field notes were recorded by one researcher (MM, a female research pharmacist) using a rapid research evaluation form designed by the research team. Recorded transcripts and text files were stored securely on trusted NHS network servers.

MAXQDA Plus 2022 (VERBI Software) was used to analyse the raw data transcripts. One author, (MM), coded the initial transcripts and summarised the findings. Cross validation of coding and analysis was independently performed by two authors, an experienced female Doctor of Pharmacy (YJ) and a male Professor of Human-Computer Interaction (EC). The final codebook was agreed by all authors. Thematic analysis was undertaken using inductive and deductive approaches, ensuring that thematic saturation was achieved, and was informed by the Technology Acceptance Model [12] and the Theoretical Domains Framework [13].

### Quantitative data analysis

We extracted diagnosis records and data on MiADE usage from the hospital’s data warehouse to compare diagnosis recording before MiADE go-live (1 November 2023 to 25 February 2024) and after MiADE go-live (26 February 2024 to 25 February 2025). Patients who opted out of data use for research and those under the care of MiADE investigators were excluded.

For inpatients under the care of participating teams, we calculated the completeness of problem lists on discharge for the first admission of each patient during the study period, considering the ICD-10 billing diagnoses to be the gold standard. We considered an ICD-10 code to be included if a SNOMED CT problem mapping to any ICD-10 code in the same block was present on or prior to the discharge date.

We assessed outpatient use of MiADE by calculating the number of encounter diagnoses and problem list entries made per encounter within a week of the clinic date, for the earliest clinic encounter per patient within the study period.

## RESULTS

Eighty-five clinicians working in a range of specialties (27 consultants, 52 resident doctors, 1 dentist, 2 general practitioners, 1 physician assistant, 1 advanced nurse practitioner and 1 midwife) and settings (10 outpatients only, 41 inpatients only, 34 both settings) consented to the study. Eight clinicians attended the post-implementation focus group, and the median duration of access to MiADE was 270 days.

Fifty-six clinicians completed the pre-MiADE questionnaire and 7 were observed in outpatient clinics, of whom 5 were re-observed post-MiADE. We also interviewed 16 patients (10 pre-MiADE, 6 post-MiADE) and 7 inpatient clinicians (see Supplementary Figure). Twelve clinicians completed the post-MiADE questionnaire.

### Clinician documentation practices

Questionnaire responses suggested that under half of clinicians (18/39) ‘always’ or ‘usually’ use problem lists in outpatient clinics, and a quarter (10/40) do so on inpatient ward rounds (Figure 2). Clinic observations corroborated this finding, with all clinicians preferentially completing documentation using free text. The freedom of choice in documentation created variation in how the information was stored, and burdened clinicians with gathering relevant data from multiple locations in a patient’s record (Focus Group; Table 1, Quotes 1-4).

**Figure 2:**
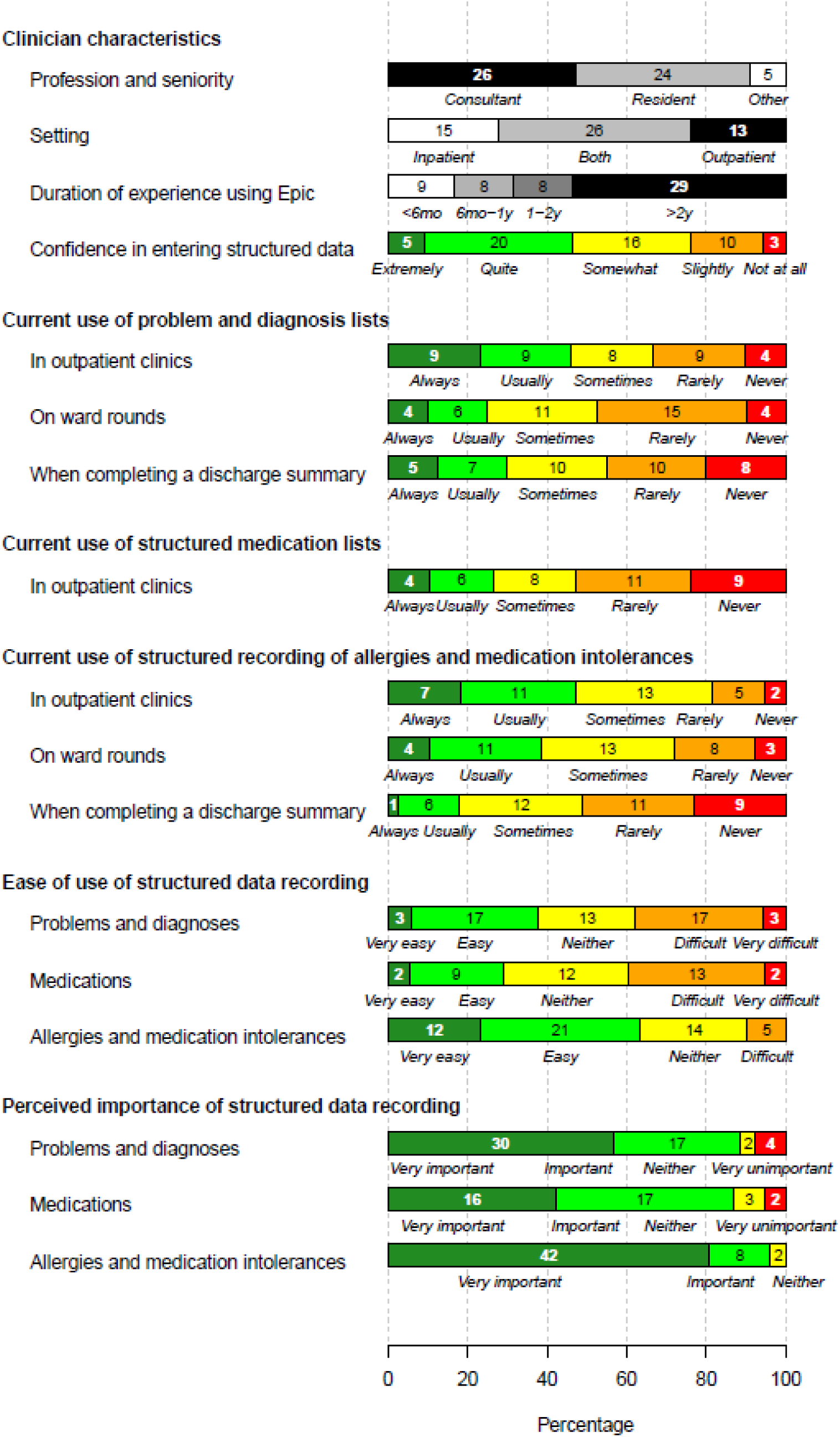
Results of pre-MiADE survey (56 responses)

**Table 1.** Quotes on baseline documentation practices in electronic health records which support themes identified in qualitative analysis.

| Quote number | Quote |
| --- | --- |
| 1 | "The amount of time every doctor spends looking at the notes to read what's wrong with the patient is enormous." (Focus Group) |
| 2 | "Lists can contain too much information which makes it difficult to read/ process quickly". (Q019, Consultant, Care of the Elderly, Pre-MiADE Survey) |
| 3 | "[Structured data]: Not generally used on ward in my experience, sometimes (particularly in elder patients) oversaturated with information, much of which is less relevant and difficult to fully digest" (Q041, FY1 Doctor, Infectious Diseases, Pre-MiADE Survey) |
| 4 | "...the ideal world ...is when you click on epic, you open a patient, you look at the problem list. It is the best place to tell you immediately what is wrong with the patient. In order to do that you need to be able to distinguish between what is your active hospital problem and what is your past medical history so those bits have to be separated off. Because if you don't understand that narrative, it's completely useless and senseless and it's just a big pile of stuff." (Focus Group) |

Clinicians also expressed confusion about when to enter structured data, whether it should be entered at initial presentation or after confirming a diagnosis (Outpatient Clinic Observations).

### Barriers and enablers of structured data entry

We identified several factors influencing structured data entry: clinicians perceptions of value, time and workload constraints, knowledge of the EHR system, team culture and the interface usability.

### Clinician beliefs

Survey data revealed that clinicians recognised the value of structured data entry (Figure 2), with over 85% of respondents rating the structured recording of problems, medications and allergies as ‘very important’ or ‘important’. Respondents mentioned numerous benefits including enhanced patient safety, improved care co-ordination, streamlined administrative processes and supporting research. However, adoption of structured data entry was inconsistent (Supplementary Table 1, Quote 5).

Structured documentation for medication and allergies was considered important and easier to enter in the EHR than problem lists (Supplementary Table 1, Quotes 6-7).

Clinicians concerns about structured documentation included data quality, maintenance, and overall usability. They voiced a lack of trust in problem lists, citing perceived inaccuracies and incompleteness. As a result, they often disregarded structured documentation and relied on free text notes, creating a negative feedback loop (Supplementary Table 1, Quotes 8-9).

Many clinicians preferred using free text to capture the nuance of patient consultations (Supplementary Table 1, Quote 10). They said free text provides “lots of opportunity to add detail” and allows them to describe how conditions “…affect the patient” (Supplementary Table 1, Quotes 11-12). Additionally, some clinicians preferred free text documentation as a permanent entry that cannot be altered by others (Supplementary Table 1, Quote 13).

Many clinicians did not perceive structured documentation as a core responsibility of their role (Supplementary Table 1, Quotes 14-15).

### Baseline knowledge and training

A shared understanding of EHR functionality and terminology is crucial to ensure quality of documentation. Some clinicians mentioned their teams organised individual sessions in the absence of central training on EHR documentation best practices (Supplementary Table 1, Quote 16).

### Workflow and time constraints

EHR documentation is often time-consuming, especially for complex or new patients. (Outpatient Clinic Observations). Clinicians reported that navigating multiple screens to review and update notes was cumbersome and inefficient, making it harder to access key patient information (Supplementary Table 1, Quotes 17-18).

Additionally, clinician workload and the demands of the clinical setting significantly impacts the time available for documentation (Supplementary Table 1, Quote 19).

### Team culture and practices

Some teams adopted strategies to encourage the use of structured data, such as templates that pull structured documentation into progress notes, providing an organised framework, and using reminders to update the problem list before discharge. In other teams, structured documentation was not prioritised, and engagement was low (Supplementary Table 1, Quote 20). Furthermore, there was recognition that inconsistent documentation practices across different teams involved in a patients care contributed to challenges with data quality (Supplementary Table 1, Quote 21).

### EHR usability

Survey responses showed a wide range of opinions regarding ease of structured data entry (Figure 2). The existing interface design required clinicians to switch between screens frequently, disrupting their workflow (Supplementary Table 1, Quotes 22-24). Clinicians also mentioned it was confusing for infrequent users as and when the system interface changed (Supplementary Table 1, Quotes 25-26).

Features that aid structured documentation, such as access via a non-intrusive side bar or reminder prompts (as for allergies and medications), were considered by some to be easier to use than the interface for entering problems (Supplementary Table 1, Quotes 27-28). Dislike of the aesthetic of structured documentation negatively impacts its use (Supplementary Table 1, Quote 29).

System usability in modifying existing entries such as medication dosages or allergy statuses, and having to search for the appropriate codes or understand the terminology for their entries also had a negative influence (Supplementary Table 1, Quotes 30-32).

### Experience using MiADE

#### Perceived usefulness

Half of survey respondents considered MiADE to be ‘very’ or ‘moderately’ useful (Figure 3). For clinicians who infrequently entered structured information, MiADE simplified the process by automating data entry and reducing manual effort (Supplementary Table 2, Quotes 33-36). Some clinicians who generally enter structured information only for their specialty commented that MiADE prompted them to add other conditions, which they usually would not do (Supplementary Table 2, Quote 37).

**Figure 3:**
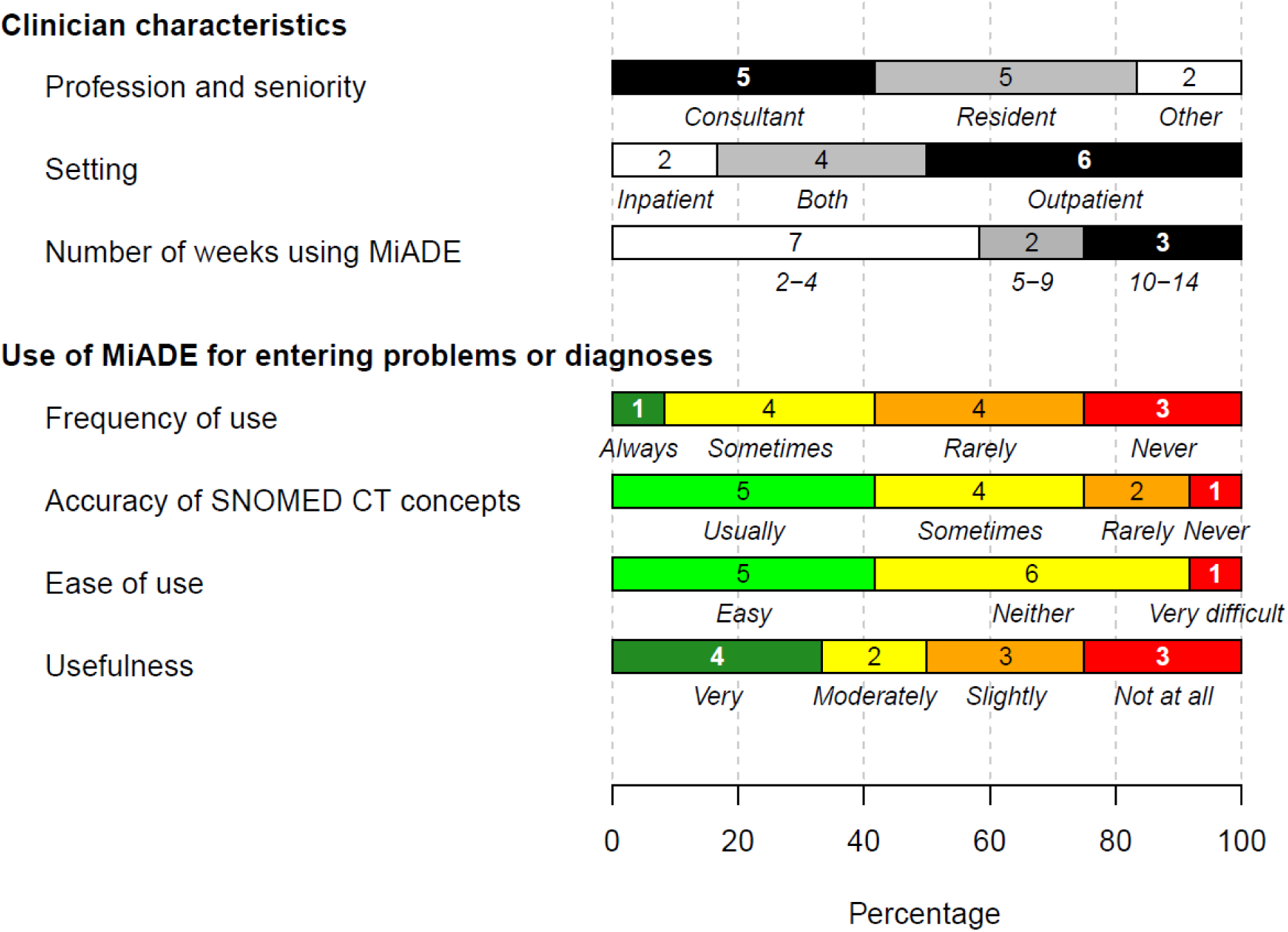
Results of post-MiADE survey (12 responses)

Despite streamlining some aspects, clinicians felt that MiADE introduced additional time to the overall documentation process (Supplementary Table 2, Quotes 38-41). and could sometimes be slow and get “stuck” (Post-MiADE Outpatient Clinic Observations).

#### Accuracy and trust

Clinicians reported that MiADE sometimes returned inaccurate suggestions or failed to recognise relevant diagnoses. These inaccuracies, particularly false positives, were reported to potentially erode clinician trust and discourage the use of MiADE (Supplementary Table 2, Quotes 42-45).

#### Engagement and training

Many clinicians did not recall undertaking the MiADE training, leading to confusion and suboptimal usage. Despite signing up to the study, some clinicians were unaware that MiADE was available to use (Field Notes, Post-MiADE Outpatient Clinic Observations; Supplementary Table 2, Quote 46).

Engagement and acceptability of MiADE was strongly linked to the clinician’s usual workflow. Those clinicians who already used structured documentation on EHR preferred to continue with that whereas those who did not enter structured information before found MiADE more beneficial (Supplementary Table 2, Quotes 47-48).

#### System improvement suggestions

Several clinicians stated more automation of coding would improve structured documentation,however only one referred to the process of actively reviewing the MiADE suggestions as an important step to minimise the risk of incorrect entries being added to the patients record (Supplementary Table 2, Quotes 49-50).

### Patient perspectives on documentation

Patients confirmed that they were satisfied with their doctor’s use of the computer during their consultation. Some patients did not notice the clinician using the computer and were more concerned with the outcome of the consultation, whereas others felt reassured that their doctor was documenting what they were saying during the consultation and liked when the clinician used the computer as an aid in the consultation.

Patients did not seem to be aware of the potential uses or benefits of structured information within the EHR, and did not notice any difference in the consultation process following the implementation of MiADE.

### Impact of MiADE on structured data recording

Forty of the 85 participating clinicians carried out at least one outpatient consultation during the study period. Among the 12,309 outpatient consultations analysed, we found an overall 36% increase in the mean number of problems or diagnoses per encounter from 0.665 to 0.904 after MiADE go-live (increase of 0.239; 95% CI 0.194, 0.284; p < 0.001 by t test) (Table 2). However, only 14 outpatient clinicians actually received MiADE suggestions during the study period. Amongst this group, there was also an increase in the proportion of encounters with at least one problem or diagnosis entered, from 49.6% to 54.0% (p < 0.001 by proportion test) (Table 2).

**Table 2.** Recording of structured diagnoses in outpatient encounters before and after MiADE go-live, by clinician’s MiADE activation status. ‘Diagnoses’ refers to encounter diagnoses or problem list entries created during or within one week of the consultation. Each patient was included only once per time period.

|  | **Outpatient clinicians actually using MiADE (n = 14)** |  | **Outpatient clinicians with MiADE available but not using (n = 26)** |  |
| --- | --- | --- | --- | --- |
| **Time period** | **Pre-MiADE** | **Post-MiADE** | **Pre-MiADE** | **Post-MiADE** |
| Number of patients | 1342 | 3408 | 2639 | 4920 |
| Median age in years | 43 | 44 | 48 | 46.5 |
| Percentage (N) female | 62.9% (844) | 59.3% (2022) | 58.4% (1541) | 56.6% (2784) |
| Total number of diagnoses | 1081 | 3525 | 1567 | 4001 |
| Number (%) of diagnoses entered using MiADE | 0 | 429 (12.2%) | 0 | 0 |
| MiADE acceptance rate with original filter (before 15 April 2024) | - | 21.6% (155/719) | - | - |
| MiADE acceptance rate with amended filter (after 15 April 2024; only diagnoses under a relevant header) | - | 77.0% (274/356) | - | - |
| Number of encounters with at least one diagnosis | 666 | 1842 | 1081 | 1972 |
| Percentage (95% CI) of encounters with at least one diagnosis | 49.6% (46.9%, 52.3%) | 54.0% (52.4%, 55.7%) | 41.0% (39.1%, 42.9%) | 40.1% (38.7%, 41.5%) |
| Mean (SD) number of diagnoses per encounter | 0.806 (1.11) | 1.03 (1.47) | 0.60 (0.95) | 0.81 (1.51) |

Among inpatient teams, there was no significant increase in the number of problems recorded after MiADE go-live, nor in the proportion of ICD-10 codes with a corresponding problem list item (Supplementary Table 4). There was a variable level of engagement with MiADE between inpatient teams, with few users consented and very little usage of problem lists in some teams (Supplementary Table 5).

The acceptance rate of MiADE suggestions improved from 22.6% (192/850) to 58.3% (309/530, p < 0.001 from proportion test) after amending the filtering of MiADE suggestions to return only diagnoses under a relevant heading. No patient safety incidents related to MiADE were reported during the study.

## DISCUSSION

### Key results

This study demonstrates the safe deployment of an NLP based structured documentation tool (MiADE) within a live clinical EHR environment. The study also highlights the challenges in engaging with clinicians to influence EHR use within their busy workload. Fewer than a third of signed up participants activated MiADE and received NLP suggestions, but nevertheless there was a 36% increase in the overall structured recording of outpatient diagnoses among patients seen by study participants. Use of MiADE varied considerably between clinicians, influenced by individual preferences, specialty specific needs, and familiarity with structured documentation.

Qualitative findings from interviews, observations and survey data demonstrate the persistent challenges in embedding structured data entry into clinical workflows. While clinicians appreciate the importance of structured documentation, there was uncertainty about when and by whom structured information should be entered, and significant baseline variation in use of structured data in the EHR.

Time constraints and workflow disruptions were consistently cited as a barrier to high-quality documentation, consistent with previous studies [14–16]. Research on EHR usability has found that systems often interrupt clinical workflows, add to clinicians’ cognitive load, and contribute to note bloat, clinician burnout, and reduced patient interaction time [17–19].

Clinicians reported that MiADE was a helpful adjunct to streamline notetaking in straightforward cases, but the SNOMED CT suggestions were sometimes inaccurate or not specific enough for specialty needs. Variation in user experience is similar to findings in other studies, where roughly half of clinicians reported positive outcomes with AI-based documentation tools while others experienced no time-saving benefit [20].

From the patient’s perspective the use of structured documentation was not a major concern. Consistent with previous studies, we found that patients prioritised communication quality and clinical outcomes over the method the clinician used to take notes [21].

### Recommendations

We suggest that buy-in from senior clinical leadership is essential for structured data recording to be considered an essential part of professional practice. Training and ongoing clinician engagement are also important to ensure that tools are as useful as possible [18,19].

### Limitations

Our study yielded novel insights into the use of point of care NLP for structured data recording, but had several limitations.

First, participation was voluntary, relying on clinicians to contribute their time. This may have introduced selection bias; those with a particular interest in EHR may have been overrepresented. Engagement among study participants was suboptimal. Sustained and longer term use would have enabled triangulation of qualitative insights with quantitative usage data.

Second, the MiADE system implementation was limited by the specific EHR (Epic) and the existing NoteReader component. We were limited in our ability to control how the MiADE results were displayed to the user. For example, we had to prefix each diagnosis label with a number in order to ensure the suggestions appeared in the same order as in the text note (Figure 1). As MiADE does not retain the analysed text, we were unable to evaluate the accuracy of suggestions provided during the study, although we have validated the NLP models previously [11].

Third, the quantitative evaluation was limited as there was no control group, so it is possible that the difference in diagnosis recording before and after MiADE go-live may have been due to factors other than MiADE.

## CONCLUSION

MiADE was successfully implemented in a live clinical EHR across a range of care settings, and was associated with an increase in diagnosis recording in outpatients. Adoption among clinical users was mixed, shaped by differences in workflow, specialty requirements, and individual perceptions of the system’s values. Future work should prioritise clinician co-design and better integration of structured documentation into clinical workflows.

## Data Availability

The study involves analysis of patient data which are not available externally in order to maintain confidentiality.

## ACKNOWLEDGEMENTS

We acknowledge the support of the UCL/UCLH NIHR Biomedical Research Centre and the staff and patients at UCLH who participated in this research.

## AUTHOR CONTRIBUTIONS

Conceptualization: LZ, FA, RD, YJ, EC, AS. Data curation: MM, JR, MA, JB, LZ, GP, FA, RD, YJ, EC, AS. Formal analysis: MM, LZ, GP, YJ, EC, AS. Funding acquisition: JB, LZ, FA, RD, YJ, EC, AS. Investigation: MM, JR, MA, JB, LZ, GP, YJ, EC, AS. Software: JB, LZ, GP. Writing – original draft: MM. Writing – review & editing: MM, JR, MA, JB, LZ, GP, FA, RD, YJ, EC, AS.

