## Supplementary Tables for "Clinical evaluation of a natural language processing system for assisting structured diagnosis recording at the point of care: MiADE (Medical Information AI Data Extractor)"

### Supplementary Table 1: Factors influencing structured data recording in EHR

| Theme | Quote number | Quote |
|---|---|---|
| | 5 | “I have only entered medications a couple of times when in clinic, but I do believe this is important and I should do it more frequently going forwards.” (Q006, Physican Associate, Clinical Pharmacology/ General Internal Medicine, Pre-MiADE Survey) |
| | 6 | “Probably most important part of record. [Structured documentation for allergies]” (Q015, Midwife, Obstetrics and Gynaecology, Pre-MiADE Survey) |
| | 7 | “This [structured medication entry] is much easier than problem lists and much more helpful and reduces drug errors.” (Q040, ST1-2 Doctor, Medicine for the Elderly, Pre-MiADE Survey) |
| | 8 | “…problem lists when you open it… there’s no confidence in it… so people ignore it and they go to the free text.” (Focus Group) |
| | 9 | “Don't use it [structured documentation] in my clinical flow as often incomplete” (500, Consultant, Cardiology, Pre-MiADE Survey) |
| *Perceptions and beliefs* | 10 | “Very important but time consuming and I enjoy the creativity of free texting my own problem list. Gives me ownership and imparts my part of critical thinking into the patients notes.” (Q040, ST1-2 Doctor, Medicine for the Elderly, Pre-MiADE Survey) |
| | 11 | “lots of opportunity to add detail” (Q004, Consultant, Respiratory, Pre-MiADE Survey) |
| | 12 | “…affects the patient” (Q002, Consultant, Surgery, Pre-MiADE Survey). |
| | 13 | “As I see patients with complex multisystemic manifestations I concentrate on creating good notes/ letter as this cannot be edited by other users.” (Q036, Consultant, Rheumatology, Pre-MiADE Survey) |
| | 14 | “…[structured medication documentation] not so relevant in my surgical practice.” (700, Consultant, Colorectal and Sarcoma, Pre-MiADE Survey) |
| | 15 | “[Structured documentation]: Not needed in my role.” (Q018, ST6-8 Doctor, Infectious Diseases, Pre-MiADE Survey) |
| *Knowledge to enter structured documentation* | 16 | “…there is a total lack of basic training in how to use (EHRS) functionality or …terminology.” (TL116, Consultant, Cardiology, Post MiADE Interview) |

| Theme | Quote number | Quote |
|---|---|---|
| *accurately* | | |
| *Time/ Workflow* | 17 | “The amount of time every doctor spends looking at the notes to read what's wrong with the patient is enormous.” (Clinician Workshop) |
| | 18 | “It is time consuming, but I find it easy to do. It is a bit clunky as you have to click between various screens so if someone has a lot of information to add for a problem or multiple problems it can be time consuming.” (Q006, Physican Associate, Clinical Pharmacology/ General Internal Medicine, Pre-MiADE Survey) |
| | 19 | “I would say time is the biggest limiting factor – during a busy ward round or large job list it is difficult to find time to update a patients problem list.” (Q022, FY1 Doctor, Medicine for the Elderly, Pre-MiADE Survey) |
| *Team* | 20 | “I forward the [MiADE study] email to my inner circle of colleagues, who by the way are not great users [of EHRS] …and no one pays attention to this… no-one." (DC1206, ST6-8 Doctor, Obstetrics and Gynaecology, Post-MiADE Interview) |
| | 21 | “It's when in an inpatient setting, when there's 20 different doctors when there's handovers from the Monday to Thursday you know no one currently believes in it so they don't use it.” (Clinician Workshop) |
| *User interface/ Usability* | 22 | “Time consuming and interrupts flow of my thought process of crafting a WR note or clerking” (Q040, ST1-2 Doctor, Medicine for the Elderly, Pre-MiADE Survey) |
| | 23 | “The reason why people free text… you can't read what's wrong with the patient and write at the same time.” (Clinician Workshop) |
| | 24 | “The 'problems' tab cannot be viewed in the right-hand panel, and therefore you cannot enter problems and have notes open at the same time. This requires lots of flicking between notes and problems which is time consuming, making me less likely to do it.” (Q039, CT1-2 Doctor, Clinical Pharmacology, Pre-MiADE Survey) |
| | 25 | “Tabs keep moving on your [EHR] screen with updates and lack of use. Don't have time to search.” (Q019, Consultant, Medicine for the Elderly, Pre-MiADE Survey). |
| | 26 | “Time consuming and unclear if the data I have added goes to the correct part” (Q019, Consultant, Medicine for the Elderly, Pre-MiADE Survey) |

| Theme | Quote number | Quote |
|---|---|---|
| | 27 | “…has its own little box on the side, less clicks required.” (300, ST6-8 Doctor, Surgery, Pre-MiADE Survey) |
| | 28 | “…very easy to find and frequently pops out (when prescribing for example)” (DC1206, ST6-8 Doctor, Obstetrics and Gynaecology, Pre-MiADE Survey) |
| | 29 | “…looks cluttered when using structured entry, end up inputting myself” (500, Consultant, Cardiology, Pre-MiADE Survey) |
| | 30 | “…where dosages require two strength tablets of the same agent …it is incredibly difficult to alter this efficiently.” (100, Consultant, Respiratory, Pre-MiADE Survey) |
| | 31 | “Can be clunky and sometimes difficult to find the right phrase/ code for the problem” (Q027, Consultant, Medicine for the Elderly, Pre-MiADE Survey) |
| | 32 | “Differentiating between inpatient problem, PMH etc not clear.” (Q016, Consultant, Infectious Disease, Pre-MiADE Survey) |

### Supplementary Table 2: Impact of MiADE on structured data recording

| Theme | Sub-theme | Quote number | Quote |
|---|---|---|---|
| *MiADE Performance* | *Utility* | 33 | “Recognises conditions and diseases and prompts me to add it/ them to problem list.” (DC1206, ST6-8 Doctor, Obstetrics and Gynaecology, Post-MiADE Survey) |
| | | 34 | “…it just makes things easier, for me anyway, …I update the problem list a lot more now than before, just because it’s easier to do it, …it’s basically …a few clicks, whereas before you’d have to go to the problem list, like look through it, you know, find what suits, click it, whatever, whereas this one already tells you what you, it kind of like detects everything that you’ve written down already in your notes so you just have to either approve or decline.” (NA1007, FY2 Doctor, Clinical Pharmacology, Post-MiADE Interview) |
| | | 35 | “Simplifies notes on ward rounds.” (400, ST6-8 Doctor, Clinical Pharmacology, Post-MiADE Survey) |
| | | 36 | “…it makes it significantly easier, it makes it less faffy, it makes you much more likely to do it and it makes it clearer than trying to think, go through the sidebar, personal history and type in you know, it just pulls, it integrates throughout into the my clinical note setup that we use and it's obviously just that one button click for note read and then you have all your diagnosis updates.” (Q011.DW0708, Consultant, Urgent Care/ Accident and Emergency, Post-MiADE Interview) |
| | | 37 | “My ENT diagnoses on a daily basis are quite few …However, I would not look for non-ENT problems in a busy outpatients clinics. MiADE lets me do that.” (300, ST6-8 Doctor, Surgery, Post-MiADE Survey) |
| | | 38 | “It's made it easier but it has added to time to my clinic encounter.” (600, Consultant, Rheumatology, Post-MiADE Survey) |
| | | 39 | “It is slow. In a busy clinic where each minute matters, I sometimes don't wait for it and enter problems they way I used to.” (300, ST6-8 Doctor, Surgery, Post-MiADE Survey) |

| Theme | Sub-theme | Quote number | Quote |
|---|---|---|---|
| | | 40 | “It slows down a bit the documentation process, when reviewing the notes.” (DC1206, ST6-8 Doctor, Obstetrics and Gynaecology, Post-MiADE Survey) |
| | | 41 | “Slowed down signing of my note while it’s processing the note itself.” (Q015, Midwife, Obstetrics and Gynaecology, Post-MiADE Survey) |
| | *Accuracy* | 42 | “It brings up problems already charted.” (600, Consultant, Rheumatology, Post-MiADE Survey) |
| | | 43 | “Some of the suggestions from MiADE don't make sense e.g. suggesting falls or covid for all patients, despite that not being listed as a problem. Also, some of the suggestions have numbers at the front e.g. 00falls and then when you go to select it, it says that SNOMED code doesn't exist” (Q006, Physican Associate, Clinical Pharmacology/ General Internal Medicine, Post-MiADE Survey) |
| | | 44 | “I think false positives at the start, is bad, I think you always wanna start with the basic less is more.” (Clinician Workshop) |
| | | 45 | “Often inaccurate or leaves out diagnoses even when they are entered in a way that should be legible to the system e.g. without blank lines.... When problems are recognised by the system, it makes coding a problem list into Epic a lot easier and faster” (SE1007, CT1-2 Doctor, Clinical Pharmacology, Post-MiADE Survey) |
| *User training and engagement* | | 46 | “I hadn't really noticed it [MiADE] but will try and engage more in future” (Q001, Consultant, Neurology, Post-MiADE Survey) |
| | | 47 | “I am a "fan" of the Problem List and keep reviewing/updating it all the time, it's usually the 1st thing I check when I open patient's notes; therefore, has not made a great difference. But I can see the potential and think will make a great difference in the documentation of those clinicians that don't regularly review or update the Problem List.” (DC1206, ST6-8 Doctor, Obstetrics and Gynaecology, Post-MiADE Survey) |
| | | 48 | “I think it is not intuitive enough yet for me to use, I prefer currently to enter the problems manually to the problem list” (Q006, Physican Associate, Clinical |

| Theme | Sub-theme | Quote number | Quote |
|---|---|---|---|
| | | | Pharmacology/ General Internal Medicine, Post-MiADE Survey) |
| System improvements for MiADE | | 49 | Re: improvements to structured documentation - “More intuitive interface. Capacity to be automatically updated as the patient’s condition progresses.” (Q043, ST3-5 Doctor, General Practitioner, Pre-MiADE Survey) |
| | | 50 | “…like its not unsafe in any way but it still has to be approved by a clinician to actually add the codes or whatever so I think it will be completely appropriate that it was used at like a wider level across the trust would be my impression.” (Q011.DW0708, Consultant, Urgent Care/ Accident and Emergency, Post-MiADE Interview) |

### Supplementary Table 3: Summary of MiADE usage by clinician setting

| | Setting(s) in which clinician works | | | |
|---|---|---|---|---|
| | **Inpatient only** | **Outpatient only** | **Both inpatient and outpatient** | **Overall / total** |
| No. of clinicians who consented to study and had MiADE available | 41 | 10 | 34 | 85 |
| No. of clinicians who actually used MiADE | 9 | 4 | 11 | 24 |
| No. of MiADE suggestions | 195 | 228 | 847 | 1270 |
| No. of MiADE suggestions accepted | 72 | 107 | 322 | 501 |
| MiADE acceptance rate with original filter (before 15 April 2024) | 28.2% (37/131) | 46.8% (102/218) | 10.6% (53/501) | 22.6% (192/850) |
| MiADE acceptance rate with amended filter (after 15 April 2024; only diagnoses under a relevant header) | 54.7% (35/64) | 50% (5/10) | 77.8% (269/346) | 73.6% (309/420) |

### Supplementary Table 4: Problem list usage pre and post MiADE go-live for participating inpatient teams

| **Time period** | **Pre-MiADE** | **Post-MiADE** |
|---|---|---|
| Number of patients (admissions) | 1263 | 3336 |
| Median age in years | 69 | 67 |
| Percentage (N) female | 49.2% (622) | 48.2% (1609) |
| Total number of hospital problems | 1371 | 2646 |
| Total number of non-hospital problems | 247 | 1247 |
| Mean (SD) number of problems per patient | 1.28 (2.12) | 1.17 (2.30) |
| ***Comparison of problem lists with ICD-10 billing diagnoses*** | | |
| Percentage (95% CI) of patients whose ICD-10 primary diagnosis is included in the problem list | 8.9% (7.4%, 10.7%) | 5.1% (4.4%, 5.9%) |
| Mean number of secondary diagnoses per patient | 14.8 | 14.4 |
| Percentage (95% CI) of ICD-10 secondary diagnoses included on the patient's problem list | 3.2% (3.0%, 3.5%) | 2.0% (1.8%, 2.1%) |

### Supplementary Table 5: Problem list usage pre and post MiADE go-live by inpatient team

| | | Number of patients (admissions) | | Mean (SD) number of problems per patient | | Percentage (95% CI) of primary ICD-10 diagnoses with corresponding problem list items | |
|---|---|---|---|---|---|---|---|
| **Team** | **Number of inpatient clinicians consented to MiADE study** | **Pre-MiADE** | **Post-MiADE** | **Pre-MiADE** | **Post-MiADE** | **Pre-MiADE** | **Post-MiADE** |
| Clinical pharmacology and internal medicine team | 14 | 195 | 663 | 2.0 (2.2) | 1.8 (2.5) | 15.4 (10.6, 21.2) | 11.0 (8.7, 13.6) |
| Respiratory team | 3 | 408 | 1081 | 0.39 (1.1) | 0.26 (0.88) | 3.2 (1.7, 5.4) | 0.7 (0.3, 1.5) |
| Infection team | 7 | 322 | 820 | 0.87 (1.5) | 1.1 (2.0) | 10.6 (7.4, 14.4) | 6.7 (5.1, 8.6) |
| Medicine for elderly team | 7 | 338 | 772 | 2.4 (2.9) | 2.0 (3.2) | 10.7 (7.6, 14.4) | 4.4 (3.1, 6.1) |
