## Supplementary figures and images for "Clinical evaluation of a natural language processing system for assisting structured diagnosis recording at the point of care: MiADE (Medical Information AI Data Extractor)"

### Supplementary Figure

**Supplementary Figure : Study timeline**

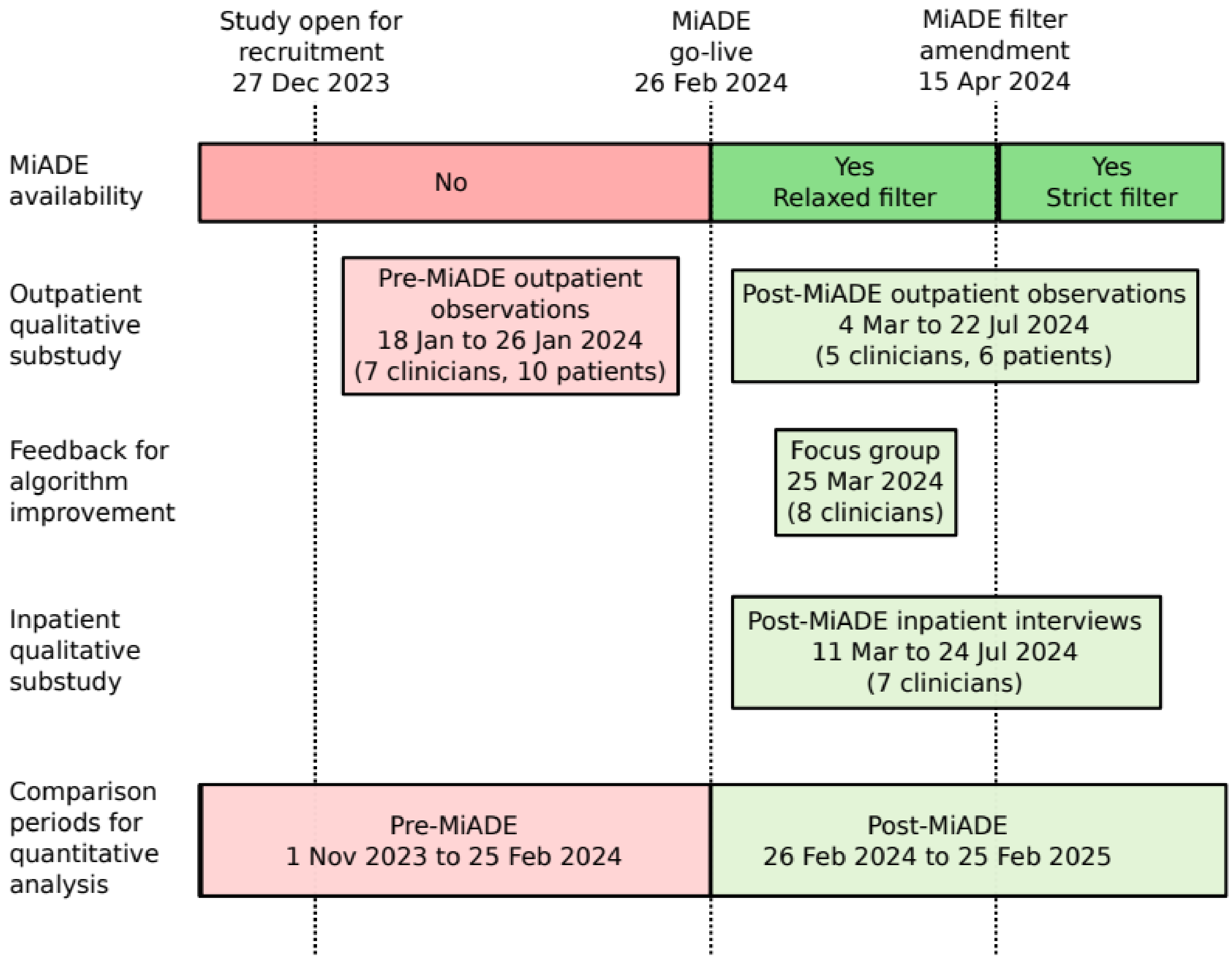
